# Dietary risk factors for monoclonal gammopathy of undetermined significance in a racially diverse population

**DOI:** 10.1101/2023.09.05.23294947

**Authors:** Janine M. Joseph, Jens Hillengass, Li Tang, Alexander M. Lesokhin, Ola Landgren, Saad Z. Usmani, Kirsten B. Moysich, Susan E. McCann, Urvi A. Shah

## Abstract

Monoclonal gammopathy of undetermined significance (MGUS) – a precursor of multiple myeloma – is associated with shorter lifespan and cardiac, renal, neurologic, and immune-related comorbidities. There is little known about modifiable risk factors for this condition. To determine if risk of MGUS is associated with dietary factors in a racially diverse population, we conducted a United States population-based case-control study from the National Health and Nutrition Examination Survey (1988-2004), which included 373 individuals with MGUS and 1,406 matched controls. Diet was characterized by one 24-hour dietary recall, with gram intake of individual foods and beverages aggregated into groups. Unconditional multivariable logistic regressions were used to model associations between intake of several food groups and MGUS, with odds ratios (OR) and 95% confidence intervals (95% CI) reported for the highest relative to the lowest quantile of intake. Daily gram intake of several food and beverage groups were significantly associated with MGUS. MGUS was inversely associated with whole-grain bread, oats, and rice (OR 0.70; 95% CI 0.48-1.00; *P*<0.05), fruits (excluding juice) and vegetables (OR 0.69; 95% CI 0.52-0.93; *P*=0.02), vegetables (OR 0.75; 95% CI 0.56-0.99; *P*<0.05), tomatoes (OR 0.72; 95% CI 0.51-1.00; *P*<0.05), and cruciferous vegetables (OR 0.44; 95% CI 0.26-0.74; *P*<0.01). Direct associations were observed for sugar-sweetened beverages (OR 1.34; 95% CI 1.00-1.78; *P*<0.05), sugar-sweetened soft drinks (OR 1.41; 95% CI 1.01-1.96; *P*=0.04), and artificially sweetened soft drinks (OR 1.55; 95% CI 1.04-2.33; *P*=0.03). Our study shows that diet is potentially a modifiable risk factor for MGUS.

## Key Points

Consumption of whole-grain bread, oats, and rice (*P*<0.05) and fruits and vegetables (*P*=0.02) was associated with reduced risk of MGUS.

Intake of sugar-(*P*=0.04) or artificially (*P*=0.03) sweetened soft drinks was associated with increased risk of MGUS.

## Introduction

Monoclonal gammopathy of undetermined significance (MGUS) is characterized by elevated abnormal serum proteins produced by clonal plasma cells. Although usually benign, MGUS progresses to multiple myeloma (MM) or a related disorder at a rate of approximately 1% per year^1^. Additionally, there is evidence that MGUS is associated with shorter lifespan^1, 2^ and cardiac, renal, neurologic, and immune-related comorbidities^3^. An estimated 3.0-3.6% of White and 5.9-8.4% of Black individuals have MGUS^4^.

Risk factors for MGUS include older age^4^, male sex^4^, Black race^4^, family history^4^, chronic antigenic stimulation^5^, infections and inflammatory conditions^6^, certain pesticides^4, 7^, other chemicals^5^, diabetes^8^ and obesity^9^. Obesity more than doubles the risk of progression from MGUS to MM^10^.

There are limited data on the association between diet and MGUS. In a population-based case-control study from Iceland, intake of fruit during adolescence and whole-wheat bread during middle age were inversely associated with MGUS^11^. There is, however, evidence that diet is associated with MM risk or survival^12^, including diets with higher inflammatory or insulinemic potential^13^, vegetarian or vegan diets (inversely)^14^, and several foods or dietary compounds^15–18^.

There is an unmet need to identify additional risk factors related to MGUS that are potentially amenable to intervention, as most risk factors for MGUS are non-modifiable or not easily modifiable. Given that most MM cases are preceded by MGUS and dietary risk factors have been associated with MM, studying the evidence for dietary risk factors for MGUS is warranted.

## Methods

### Study Population

The National Health and Nutrition Examination Survey (NHANES) program is administered by the National Center for Health Statistics within the Centers for Disease Control and Prevention. The goal of the program is to assess the health and nutritional status of the United States population. To obtain reliable estimates of subgroups, certain populations, such as older persons, African Americans, Mexican Americans, and low-income persons are oversampled. Further details are available on the Centers for Disease Control and Prevention website^19^.

### Study Sample

Data from this population-based, case-control study were obtained from four releases of NHANES (NHANES III, 1999-2000, 2001-2002, 2003-2004), which included 77,772 individuals^20^. The overall dataset was restricted to 25,095 individuals whose sera were included in the surplus sera studies of MGUS released in 2012 for participants ≥ 50 years of age from NHANES III (n=6,557), 1999-2000 (n=1,864), 2001-2002 (n=2,130), and 2003-2004 (n=2,164), and in 2015 for participants < 50 years of age from NHANES III (n=12,380) (Figure 1), as described previously^21, 22^.

**Figure 1.**
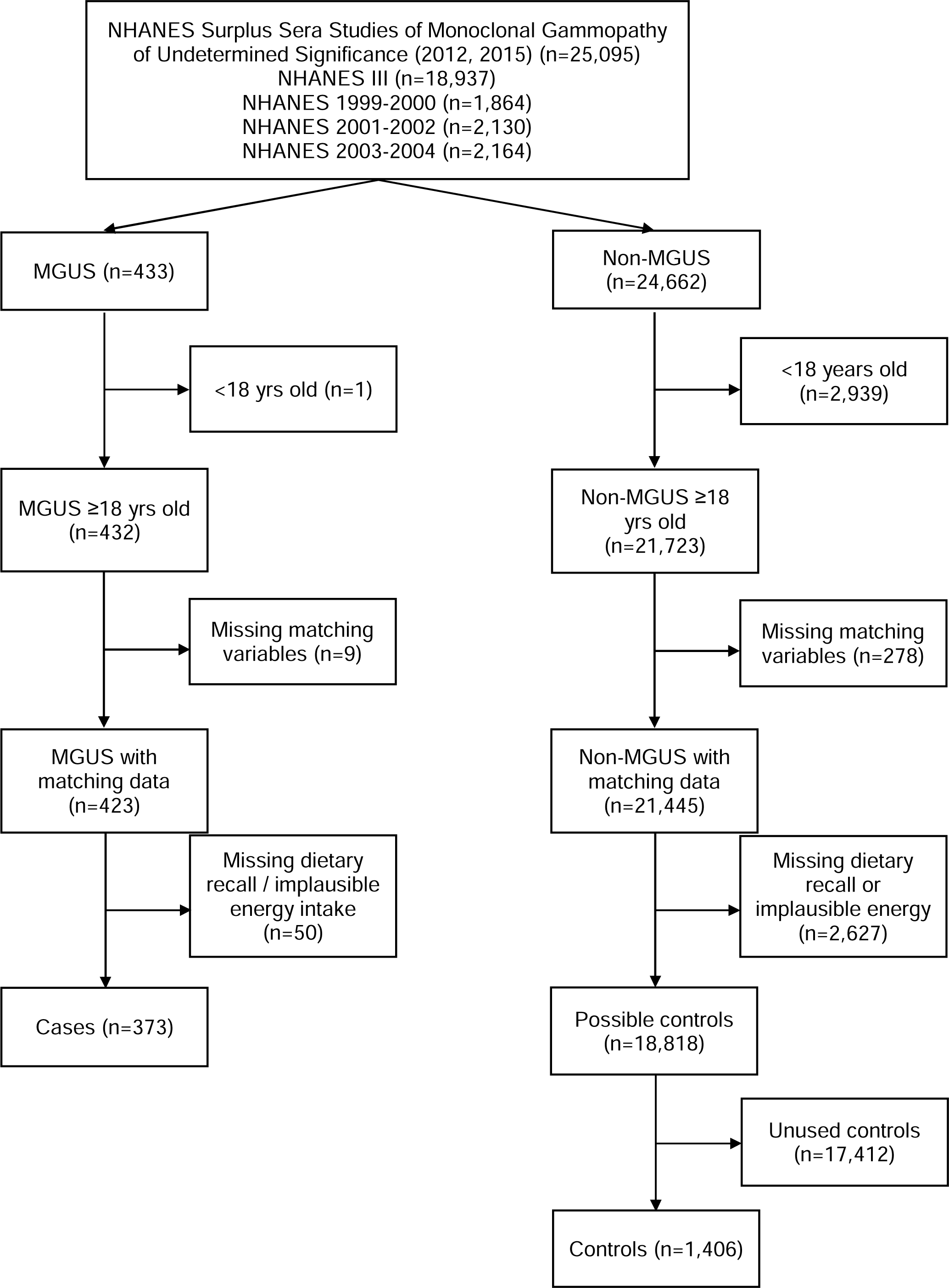
Flow chart of participants included in the National Health and Nutrition Examination Survey (NHANES) case-control study of dietary characteristics and monoclonal gammopathy of undetermined significance (MGUS)

### Case and Control Definitions

Potential cases (n=433) were defined as individuals with MGUS by established criteria^23–26^. Potential controls (n=24,662) were individuals whose sera did not meet criteria for MGUS. We excluded individuals younger than age 18 (1 case and 2,939 controls), missing matching characteristics (9 cases and 278 controls), missing 24-hour dietary recall (DR), or implausible total energy intake (<500 or >3500 kilocalories per day [kcal/d)^27^ (50 cases and 2,627 controls), leaving a total of 373 cases and 18,818 possible controls. Controls were frequency matched at a rate of four controls to one case on five criteria: NHANES release, sex, race/ethnicity, age at interview (five-year categories), and body mass index (BMI) category (<18.5, 18.5-24.9, 25.0-29.9, and ≥30.0 kg/m^2^) (Figure 1).

### Descriptive Characteristics and Dietary Measures

Descriptive characteristics were available through NHANES interview questionnaires done prior to MGUS ascertainment^28^. BMI was obtained by physical examination. Individuals with MGUS were risk stratified based on International Myeloma Working Group criteria (non-IgG-type MGUS, M protein concentration >1.5 g/dL, and/or abnormal serum free light chain ratio) into low-risk MGUS (LRMGUS, zero risk factors) and intermediate/high-risk MGUS (IHRMGUS, one or more risk factors)^29^.

Dietary practices were assessed with a DR that captured food and beverage intake for 24 hours through midnight prior to the interview. Based on the DR, NHANES provides individual estimates of total energy intake (kcal/d), macronutrients (g/d), and micronutrients. Additionally, the DR provided detail on individual foods consumed, classified by their eight-digit food code assigned by the United States Department of Agriculture (USDA). The first digit identifies the food group, as described in the USDA Food and Nutrient Database for Dietary Studies documentation^30^: (1) milk and milk products; (2) meat, poultry, fish, and mixtures; (3) eggs; (4) legumes, nuts, and seeds; (5) grain products; (6) fruits; (7) vegetables; (8) fats, oils, and salad dressings; and (9) sugars, sweets, and beverages. We divided (6) fruits into (6a) fruits (excluding juices) and (6b) fruit juices, and we created a combined “fruits (excluding juice) and vegetables” group.

Subsequent digits of the USDA food code allow for foods to be assigned to ever more specific groups. We created 12 more custom food groups based on the second or third digit or, in a few instances, by selection of individual foods based on their eight-digit code. These custom groups provided insights into the drivers of an association for a primary food group or because of prior associations with MGUS or cancer (Supplementary Table S1). For each primary and custom food group, we calculated each participant’s total g/d consumed. Consumption of each food group (g/d) was divided into tertiles (T) or, for highly skewed distributions, quantiles (Q), based on the distribution in the controls. Where more than 90% of the controls had no intake, the sample was divided into “None” or “Any” consumption.

The Healthy Eating Index (HEI), a measure of diet quality used to assess how well a set of foods aligns with the Dietary Guidelines for Americans, was available for NHANES III and calculated from DR data. Additionally, NHANES III administered an extensive FFQ which was included as supplementary analyses for this study. This queried monthly intake of 60 items. Each item was mapped to 11 primary and 10 custom FFQ food groups, which largely aligned with the USDA-based DR food groups described above (Supplementary Tables S2 and S3). Monthly consumption (times/month) was assigned into tertiles or quantiles, based on the distribution in controls.

### Statistical Methods

Key demographic and health-related characteristics of cases and controls were compared using Student’s t-tests for differences in means or chi-squared tests for differences in proportion.

Due to the frequency-match between cases and controls, i.e., not unique matching, unconditional multivariable logistic regression analyses were used to model the association between MGUS, LRMGUS, and IHRMGUS and daily intake of primary and custom food groups, with odds ratios (OR) and 95% confidence intervals (CI) as the measures of association. Firth’s corrections were applied to logistic regressions with referent groups representing ≥70% of the controls. Total daily energy intake (kcal), age in years, sex, race/ethnicity, and continuous BMI, were included as covariates in all multivariable models. Variables associated with socioeconomic status (education and family income) were not included in the final models because they did not substantially impact the modeled measures of association. P-values for food groups with more than two quantiles of intake are stated as p for trend (*P*trend) between quantile midpoints.

As secondary analyses, the association between MGUS, LRMGUS, and IHRMGUS and monthly intake of primary and custom FFQ food groups for 1,079 NHANES III participants was examined and modeled similar to the primary analyses.

In all models, a value of *P* ≤ 0.05 was considered significant. All analyses were performed using SAS 9.4 for Windows (SAS Institute, Cary, NC).

### Data Availability

Original datasets are publicly available from the Centers for Disease Control and Prevention. The datasets generated during the current study are available from the corresponding author on reasonable request.

## Results

### Participant Characteristics

There are 1,779 total participants – 373 MGUS cases and 1,406 matched controls – included in the analysis. The mean age at interview is 67.3 years. The participants are predominantly overweight/obese (69.7%), from the NHANES III cycle (61.0%), male (57.6%), non-Hispanic White (55.0%), with no college education (69.5%), married (61.0%; data not shown), have a family income above $20,000/year (52.1%) and are not current smokers (82.8%). The study was racially diverse, with 45.0% minorities, including 27.2% non-Hispanic Blacks and 16.0% Mexican Americans. There are no statistically significant differences in these characteristics between cases and controls (Table 1).

**Table 1.** Descriptive characteristics of 1,779 participants in the case-control study of dietary characteristics and monoclonal gammopathy of undetermined significance (MGUS) in the National Health and Nutrition Examination Survey (NHANES)

|  | **MGUS**<br>**(n = 373)** | **Non-MGUS<sup>1</sup>**<br>**(n = 1,406)** | **p-value<sup>2</sup>** |
| --- | --- | --- | --- |
|  | **Mean (SD)** |  |  |
| Age at interview (years) | 67.6 (14.0) | 67.2 (14.3) | 0.59 |
|  | **n (%)†** |  |  |
| BMI (kg/m<sup>2</sup>) |  |  |  |
| Underweight (<18.5) | 9 (2.4) | 17 (1.2) | 0.39 |
| Normal weight (18.5-24.99) | 106 (28.4) | 407 (29.0) |  |
| Overweight (25.0-29.99) | 151 (40.5) | 579 (41.2) |  |
| Obese (≥30.0) | 107 (28.7) | 403 (28.7) |  |
| NHANES Cycle |  |  |  |
| NHANES III | 222 (59.5) | 857 (61.0) | 0.96 |
| NHANES 1999-2000 | 38 (10.2) | 141 (10.0) |  |
| NHANES 2001-2002 | 54 (14.5) | 191 (13.6) |  |
| NHANES 2003-2004 | 59 (15.8) | 217 (15.4) |  |
| Sex |  |  |  |
| Male | 213 (57.1) | 812 (57.8) | 0.82 |
| Female | 160 (42.9) | 594 (42.3) |  |
| Race/Ethnicity |  |  |  |
| Non-Hispanic White | 197 (52.8) | 781 (55.6) | 0.13 |
| Non-Hispanic Black | 105 (28.2) | 379 (27.0) |  |
| Mexican American | 59 (15.8) | 225 (16.0) |  |
| Other Hispanic | 2 (0.5) | 1 (0.1) |  |
| Other or Multiracial | 10 (2.7) | 20 (1.4) |  |
| Education |  |  |  |
| <9<sup>th</sup> grade | 90 (24.1) | 388 (27.6) | 0.28 |
| Some high school | 78 (20.9) | 245 (17.4) |  |
| High school | 98 (26.3) | 340 (24.2) |  |
| Some college (incl. Associate's) | 54 (14.5) | 241 (17.1) |  |
| Bachelor or above | 50 (13.4) | 186 (13.2) |  |
| Family income |  |  |  |
| <\$20,000/year | 164 (44.0) | 652 (46.6) | 0.48 |
| ≥\$20,000/year | 199 (53.4) | 728 (52.0) | |
| Smoking Status |  |  |  |
| Never | 161 (43.2) | 612 (43.5) | 0.66 |
| Former | 142 (38.1) | 556 (39.5) |  |
| Current | 70 (18.8) | 236 (16.8) |  |

†Total categorical values may not sum to 100% due to missing data and rounding. MGUS=monoclonal gammopathy of undetermined significance; SD = standard deviation; N = number; <sup>1</sup> Controls and cases matched at a rate of four controls to one case on five criteria: NHANES release, sex, race/ethnicity, five-year age category, and BMI category. <sup>2</sup> Tested by Student's t-test for difference in mean or chi-squared test of differences in proportion.

### Diet and MGUS Association

#### Food Groups (Dietary Recall)

Participants report eating 1,730 kcal/d on average. Participants from NHANES III have a mean HEI score of 64, indicating an average diet that needs improvement by USDA Center for Nutrition Policy and Promotion standards^31^. This HEI score is roughly comparable to the United States national average in 1994 among individuals aged 51 and over – 64.0 for males and 67.1 for females^32^. There are no significant differences in total energy, HEI, or macronutrient intake between cases and controls (data not shown).

Multivariable models of the association between daily food intake and MGUS, LRMGUS, and IHRMGUS as defined by USDA primary (Table 2) and custom food groups (Table 3) are shown. Daily intakes of certain plant-based foods are inversely associated with MGUS. Intake of whole-wheat bread, whole oats, and brown rice shows an inverse association with MGUS (Q2 OR 0.70; 95% CI 0.48-1.00; *P*<0.05) and LRMGUS (Q2 OR 0.51; 95% CI 0.28-0.93; *P=*0.03). Intake of fruits (excluding juice) and vegetables shows a dose-dependent (*P*trend 0.02) inverse association with MGUS (T3 OR 0.69; 95% CI 0.52-0.93; *P*=0.02). Taken alone, intake of fruits (excluding juice) is inversely associated with LRMGUS (Q3 OR 0.62; 95% CI 0.38-0.99; *P*<0.05). Intake of vegetables is inversely associated with MGUS (T3 OR 0.75; 95% CI 0.56-0.99; *P*<0.05). Intake of tomatoes shows an inverse association with MGUS (Q2 OR 0.72; 95% CI 0.51-1.00; *P*<0.05). Intake of cruciferous vegetables shows an inverse association with both MGUS (Q2 OR 0.44; 95% CI 0.26-0.74; *P*<0.01) and IHRMGUS (Q2 OR 0.22; 95% CI 0.09-0.57; *P*<0.01).

**Table 2.**
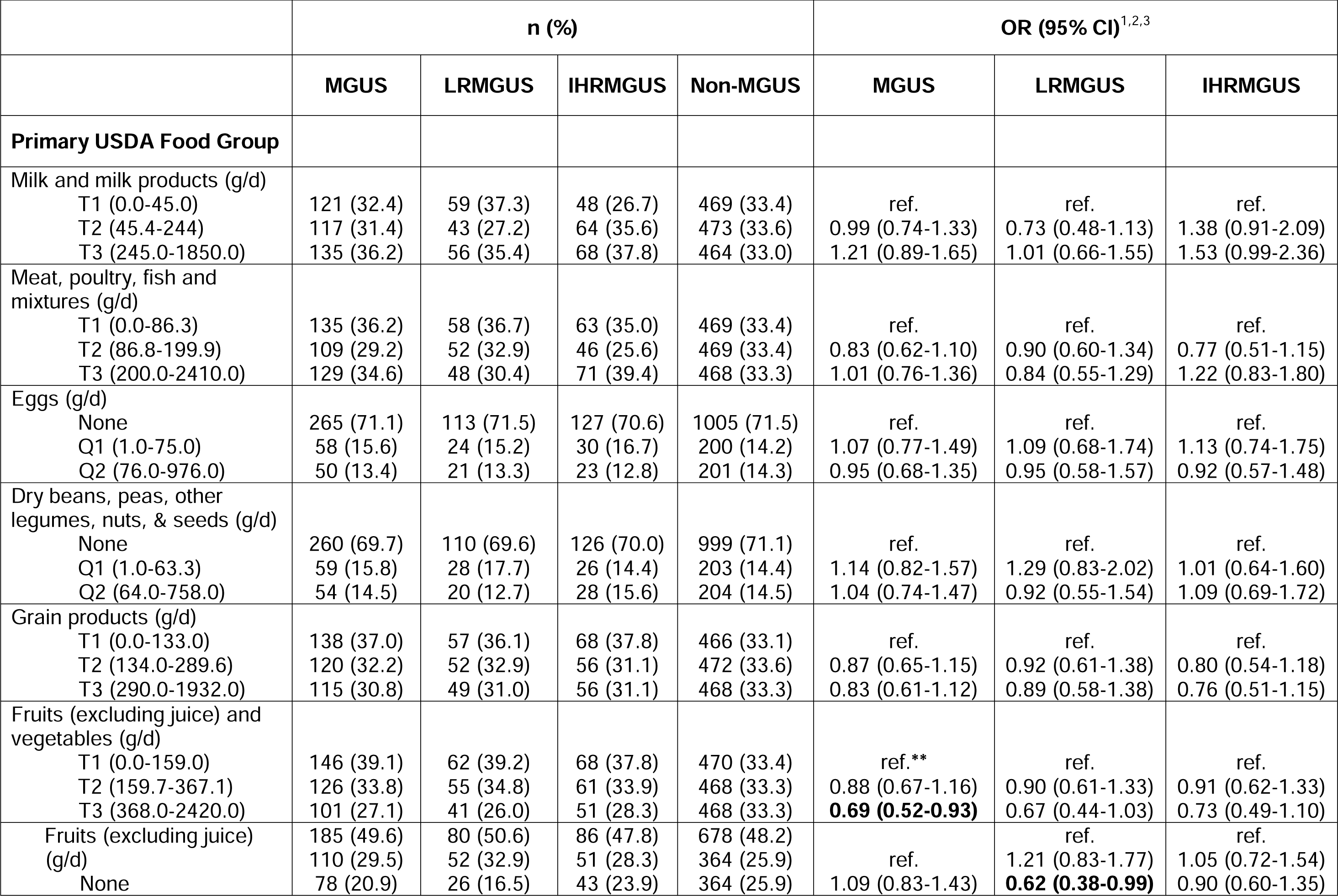
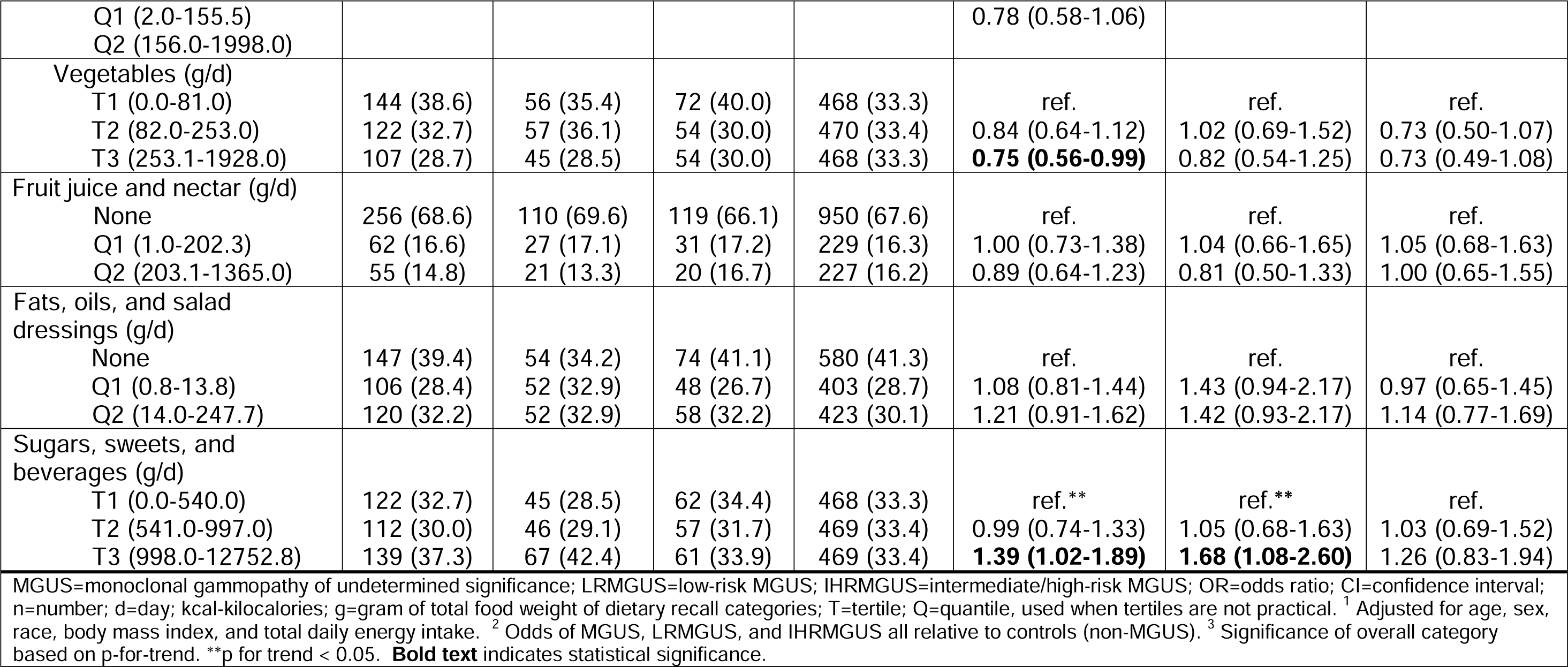
Association between daily food intake, estimated from 24-hour dietary recall data, by primary USDA food group, and overall, low-risk, and intermediate/high-risk MGUS in 1,779 participants in the case-control study of dietary characteristics and monoclonal gammopathy of undetermined significance (MGUS) in the National Health and Nutrition Examination Survey (NHANES)

**Table 3.**
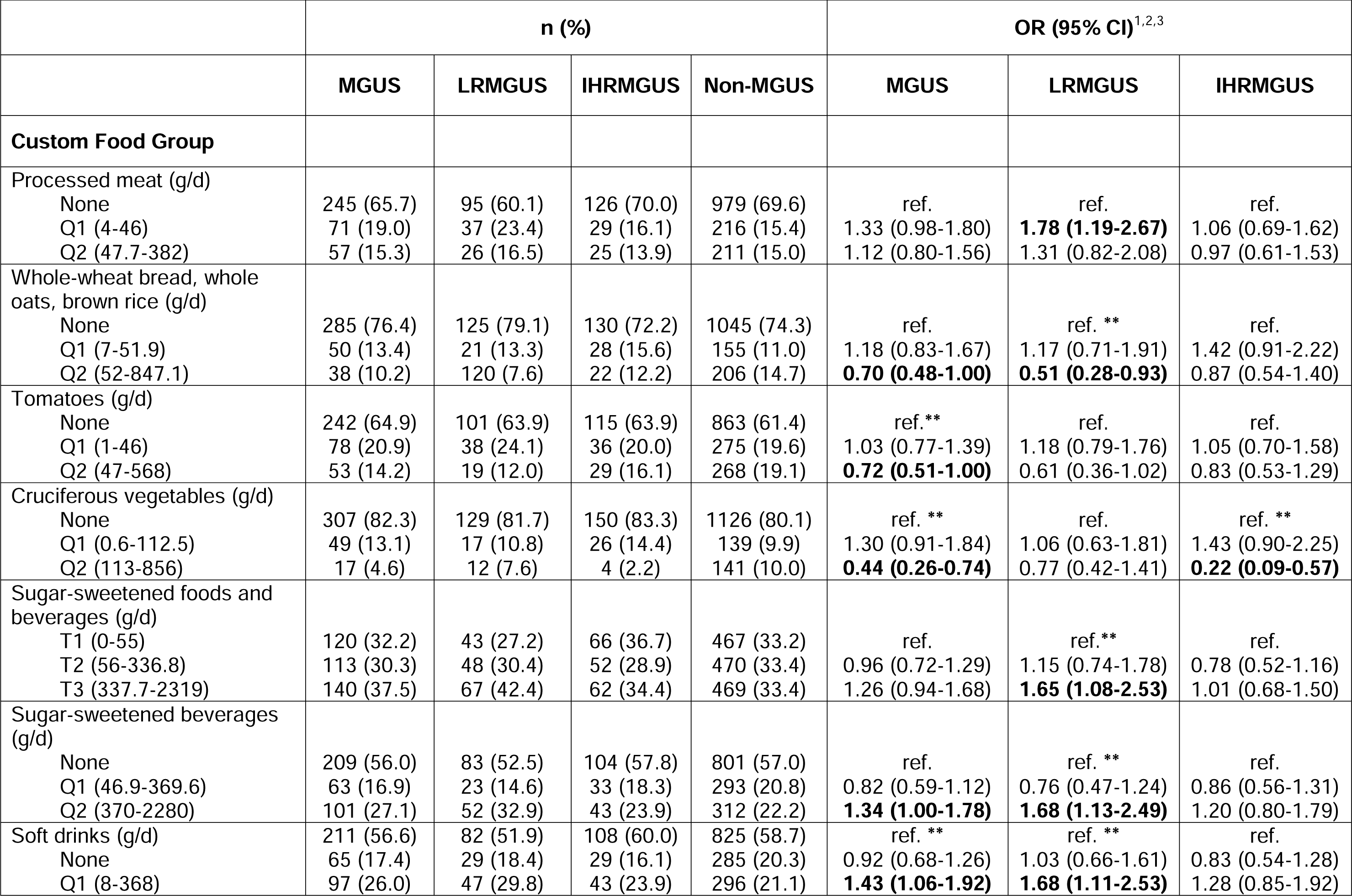
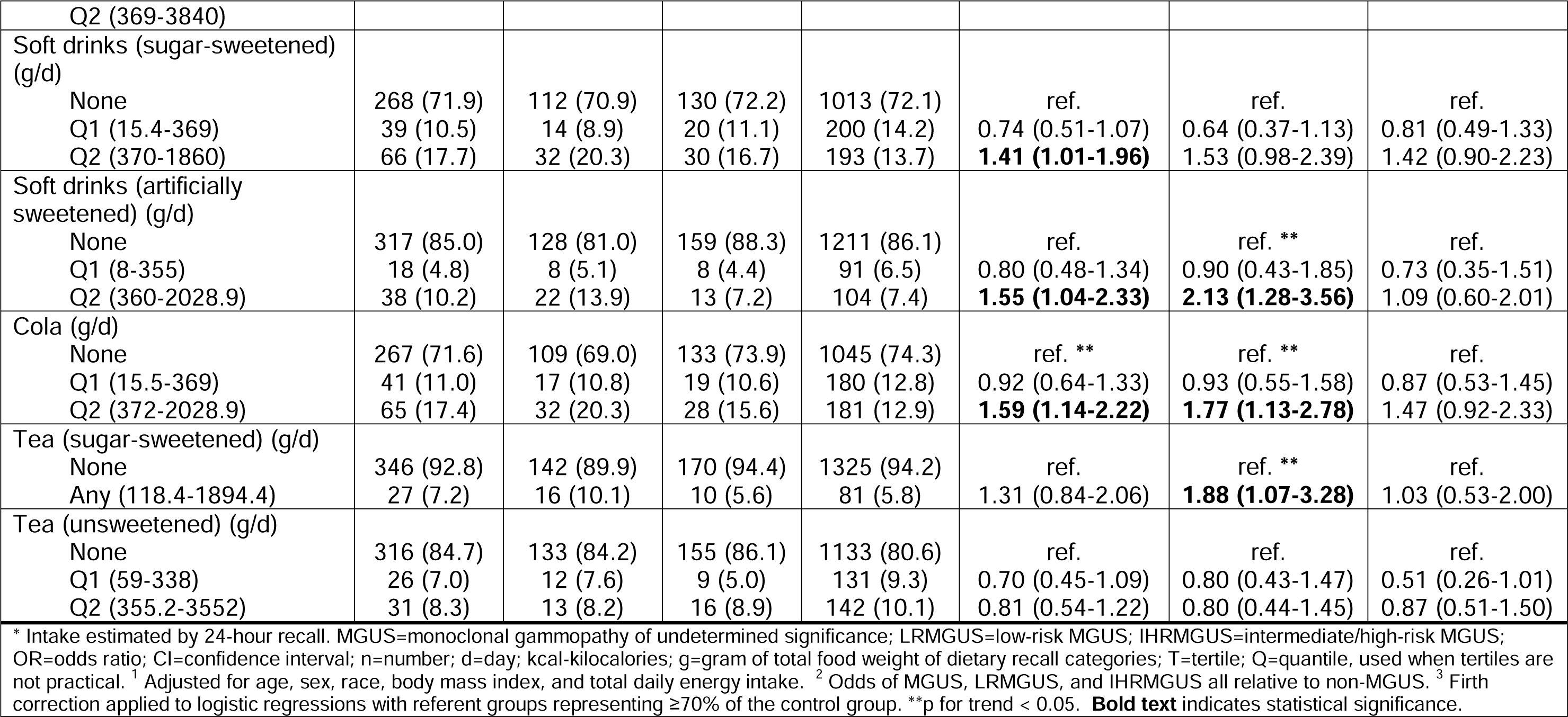
Association between daily food intake, estimated from 24-hour dietary recall data, by select custom food groups, and overall, low-risk, and intermediate/high-risk MGUS in 1,779 participants in the case-control study of dietary characteristics and monoclonal gammopathy of undetermined significance (MGUS) in the National Health and Nutrition Examination Survey (NHANES)

Daily gram intake of sugars, sweets, and beverages is associated with MGUS (T3 OR 1.39; 95% CI 1.02-1.89; *P*=0.04) and LRMGUS (T3 OR 1.68; 95% CI 1.08-2.60; *P*=0.02). This overall association is attributed to sugar- and artificially sweetened soft drinks, as described below, with no associations seen for the other major contributors to this primary USDA food group, such as desserts, candy, coffee, or alcoholic beverages (data not shown).

Intake of sugar-sweetened beverages is associated with both MGUS (Q2 OR 1.34; 95% CI 1.00-1.78; *P*<0.05) and LRMGUS (Q2 OR 1.68; 95% CI 1.13-2.49; *P*=0.01) but not with IHRMGUS. Intake of soft drinks, which includes both sugar-sweetened and artificially sweetened, is associated with MGUS (Q2 OR 1.43; 95% CI 1.06-1.92; *P*=0.02) and LRMGUS (Q2 OR 1.68; 95% CI 1.11-2.53; *P*=0.01). Similarly, intake of soft drinks (sugar-sweetened) is associated with MGUS (Q2 OR 1.41; 95% CI 1.01-1.96; *P*=0.04). There are even more pronounced associations between intake of soft drinks (artificially sweetened) and MGUS (Q2 OR 1.55; 95% CI 1.04-2.33; *P*=0.03) and LRMGUS (Q2 OR 2.13; 95% CI 1.28-3.56; *P*<0.01). Similarly, daily intake of cola (regular and diet) is associated with MGUS (Q2 OR 1.59; 95% CI 1.14-2.22; *P*<0.01) and LRMGUS (Q2 OR 1.77; 95% CI 1.13-2.78; *P*=0.01) but not IHRMGUS. Lastly, any daily intake of any tea (sugar-sweetened) is associated with LRMGUS (OR 1.88; 95% CI 1.07-3.28; *P*=0.03). By comparison, tea (unsweetened) exhibits non-significant inverse associations for MGUS, LRMGUS, and IHRMGUS, suggesting that added sugar is relevant to MGUS.

Among animal products, there is a suggestive dose-dependent (*P*trend=0.08) positive association between consumption of milk and milk products and IHRMGUS (T3 OR 1.53, 95% CI 0.99-2.36; *P*=0.05). Processed meat intake at intermediate levels is associated with LRMGUS (Q1 OR 1.78; 95% CI 1.19-2.67; *P*<0.01), but there is a non-significant association at the highest level (Q2 OR 1.31; 95% CI 0.82-2.08; *P*=0.26).

#### Usual Intake (Food Frequency Questionnaire)

Our secondary analyses of the association in NHANES III between usual monthly frequency of intake of 60 items aggregated into primary (Supplementary Table S4) and custom (Supplementary Table S5) food groups and MGUS, LRMGUS, and IHRMGUS provides support for some of the associations seen in the primary analysis of daily intake. There are dose-dependent associations between fruit juice and MGUS and IHRMGUS, and between meat, poultry, seafood, stews and MGUS. There is also an association between soft drinks (artificially sweetened) and MGUS and a borderline association with LRMGUS. Lastly, monthly intake of tea is inversely related to LRMGUS.

## Discussion

In the current study, there are associations between MGUS and daily intake of certain food groups, suggesting that lower diet quality is associated with MGUS. Notably, low daily intake of whole-wheat bread, whole oats, and brown rice, fruits (excluding juices) and vegetables, tomatoes, and cruciferous vegetables, and high daily intake of sugars, sweets, and beverages, specifically sugar- and artificially sweetened beverages are associated with MGUS. Most findings are consistent with the American Institute of Cancer Research and World Cancer Research Fund cancer prevention recommendations that recommend eating a diet rich in whole grains, vegetables, fruits, and beans and limiting consumption of sugar-sweetened drinks, fast and processed foods, and red and processed meat.

The association between MGUS and high daily intake of sugar- and artificially sweetened beverages could be due to multiple underlying mechanisms. They are a concentrated source of calories without other nutrients, such as fiber, to mitigate these effects and thus contribute to obesity, which is linked with MGUS^9^. Obesity can promote systemic inflammation and lower production of adiponectin^33^, an adipokine with anti-inflammatory, insulin-sensitizing, and anti-tumor effects, including in MM cells^34^. High intake of sugar is linked to chronic inflammation^35^ which can facilitate tumorigenesis^36^. Sugar intake can also lead to increased insulin and insulin-like growth factor 1 (IGF-1), a potent mitogen in myeloma cells^37^. Soft drinks include phosphoric acid, which may dysregulate the calcium/phosphorus balance, leading to decreased bone density and fractures^38^ and possibly tumorigenesis^39^. Sugary drinks and/or fruit juices have been previously associated with risk of overall, breast, biliary tract, colorectal, prostate, thyroid, skin, and pancreatic cancers^40, 41^.

On the contrary, intakes of whole grains, fruits, and vegetables show inverse associations with risk of MGUS, LRMGUS, and/or IHRMGUS, with a particularly strong inverse association between cruciferous vegetables and IHRMGUS. This is consistent with large epidemiologic studies that have shown a reduced cancer risk associated with intake of fruits and vegetables, especially cruciferous and green-yellow vegetables^42^, and whole grains^43^. Plant-based foods reduce cancer risk through multiple mechanisms. They are a rich source of dietary flavonoids and fiber, which decrease levels of insulin, IGF-1, and inflammation^44^. Dietary flavonoids and fiber also have beneficial effects on the immune system and microbiome, through increased levels of butyrate producers^44, 45^. Higher stool butyrate producers and butyrate concentrations have been associated with sustained minimal residual disease negativity in MM patients^46^.

Cruciferous vegetables (a group of vegetables from the *Brassica* genus of plants^47^) are unique dietary sources of glucosinolates, which hydrolyze *in vivo* toisothiocyanates (e.g. sulforaphane) and indoles, with known anti-cancer properties^48^. Tomatoes are a major source of carotenoids such as lycopene and beta-carotene that have anti-cancer properties^49^ and are a good source of vitamin C^50^. Thus, it is important to differentiate refined carbohydrates, such as those found in sugary drinks, from complex carbohydrates, such as found in whole grains, as they have opposite associations with MGUS risk and risk of other cancers.

The direct association between MGUS and intake of processed meat is consistent with its classification by the International Agency for Research on Cancer as a group 1 carcinogen, although this classification was based on associations with colorectal and gastric cancer^51^.

Associations between dietary intake and MGUS differ by risk of progression, with no food groups or nutrients significant in both LRMGUS and IHRMGUS, possibly due to the small subgroup sample sizes. Most notably, the associations between sugar- and artificially sweetened beverages and LRMGUS, the most marked associations observed in this study, are null in IHRMGUS, suggesting that there could be etiologic differences between LRMGUS and IHRMGUS; the etiology of IHRMGUS could be overwhelmed by genetics or environmental exposure, obscuring associations between diet and the condition.

Strengths of the study include the comprehensive dietary assessment, employing both quantitative (DR) and non-quantitative (FFQ) tools, capturing dietary practices in a racially diverse cohort that included minority populations. The large number of subjects without MGUS allowed us to match on multiple characteristics associated with MGUS. Blood samples were randomly evaluated for MGUS without any pre-existing suspicion of MGUS, resulting in an unbiased method used to ascertain case-control status. All participants were subject to the dietary evaluations prior to their blood being tested for MGUS, reducing the risk of recall bias.

A weakness is that certain food groups combine heterogenous foods and beverages with possibly different physiologic effects. This may have obscured associations that would have been more apparent if it were possible to analyze intake of individual food items in relation to MGUS, although that would be problematic from a statistical power standpoint. Additionally, the representation by both solid foods and beverages in certain categories complicate the interpretation, as the total gram weight consumption of the category is largely driven by intake of beverages, which tend to be consumed in larger quantities (by gram weight) and are highly variable from person to person.

Another limitation is that the assessment tools (DR and FFQ) provide a snapshot of a person’s diet which might not be connected to the biologically relevant exposure window. Longer-term dietary data, such as those used in the Iceland study^11^, would benefit future analyses.

Additionally, this study did not assess overall dietary patterns in relation to MGUS and can be addressed in future studies. Furthermore, the study population had a low-quality diet overall, with the majority of the population being below the recommended daily allowances of many micronutrients studied. This limited distribution made it difficult to detect an association between healthful dietary intakes and MGUS.

### Conclusion

This is the most comprehensive study of dietary risk factors in MGUS in a racially diverse United States population. This study provides evidence to support that diet may be a risk factor for MGUS. Given the case-control study design, it is not possible to determine a causal relationship between diet and MGUS. Future studies should focus on dietary patterns and examine dietary composition at multiple timepoints throughout life to interrogate the biologically relevant exposure window. There is also a need to examine whether the observed associations differ by race, sex, or obesity status – established risk factors for MGUS^4, 9^ –and by physical activity – an emerging risk factor for several cancers, including MM^52^. Nevertheless, the current study provides insight into how diet, a modifiable risk factor, could be related to MGUS, a condition for which very little is known regarding risk-reduction strategies.

## Acknowledgements

This study is funded in part through the NIH/NCI Cancer Center Support Grant at Memorial Sloan Kettering P30 CA008748. U.A. Shah received research and salary support from the NCI MSK Paul Calabresi Career Development Award for Clinical Oncology K12 CA184746 and American Society of Hematology Scholar Award. She also received research funding from the Paula and Rodger Riney Foundation and HealthTree Foundation. J.M. Joseph is partially supported by the Roswell Park Alliance Foundation. The authors would like to acknowledge Jeanne A. Joseph for her careful editing of the manuscript.

## Author Contributions

J.M.J., S.M. and U.A.S. designed the study and initiated this work; J.M.J. developed the statistical analysis plans and analyzed the data; J.M.J. and U.A.S. wrote the manuscript; and all authors made substantial contributions to design of the study, interpretation of results, revised the article critically and gave final approval of the manuscript to be submitted.

