## Supplemental Data for "Dietary risk factors for monoclonal gammopathy of undetermined significance in a racially diverse population"

**Supplementary Table S1.** Custom food groups constructed from NHANES 24-hour dietary recall (DR) responses in 1,779 participants in the case-control study of dietary characteristics and monoclonal gammopathy of undetermined significance in the National Health and Nutrition Examination Survey (NHANES)

| Custom DR Food Group | Major Foods in Group |
|---|---|
| Processed meat | Frankfurters / hot dogs |
| | Sausages |
| | Lunchmeats |
| Whole-wheat bread, whole oats, brown rice | Bread, whole wheat |
| | Bread, cracked wheat |
| | Oatmeal, regular |
| | Rice, brown |
| | Rice, wild |
| Tomatoes | Tomatoes |
| | Tomato sauces |
| | Tomato soups |
| Cruciferous vegetables | Greens (collard, mustard, turnip) |
| | Broccoli |
| | Cauliflower |
| | Cabbage |
| | Brussels sprouts |
| | Turnip |
| Sugar-sweetened foods & beverages | Cakes, cookies, pies, pastries, sweet crackers |
| | Sugars, sweets, candy |
| | Milk desserts, frozen |
| | Puddings, custards, and other milk desserts |
| | Sweet rolls & coffee cakes |
| | Soft drinks, sugar-sweetened (caffeinated, decaffeinated) |
| | Fruitades and drinks |
| | Tea, sugar-sweetened |

| | |
|---|---|
| | Beverages, sweetened grain drinks (oatmeal, horchata) |
| | Fruit-flavored drink, made from powdered mix |
| Sugar-sweetened beverages | Soft drinks, sugar-sweetened (caffeinated, decaffeinated) |
| | Fruitades and drinks |
| | Tea, sugar-sweetened |
| | Beverages, sweetened grain drinks (oatmeal, horchata) |
| | Fruit-flavored drink, made from powdered mix |
| Soft drinks | Soft drinks, cola types (sugar-sweetened, artificially sweetened, caffeinated, decaffeinated) |
| | Soft drinks, fruit-flavored (sugar-sweetened, artificially sweetened, caffeinated, decaffeinated) |
| | Soft drinks, other flavored (sugar-sweetened, artificially sweetened, caffeinated, decaffeinated) |
| | Water, carbonated (sugar-sweetened, artificially sweetened, unsweetened) |
| | Juice drinks, carbonated |
| Soft drinks (sugar-sweetened) | Soft drinks, cola types (sugar-sweetened, caffeinated, decaffeinated) |
| | Soft drinks, fruit-flavored (sugar-sweetened, caffeinated, decaffeinated) |
| | Soft drinks, other flavored (sugar-sweetened, caffeinated, decaffeinated) |
| | Water, carbonated (sugar-sweetened) |
| | Juice drinks, carbonated |
| Soft drinks (artificially sweetened) | Soft drinks, cola types (artificially sweetened, caffeinated, decaffeinated) |
| | Soft drinks, fruit-flavored (artificially sweetened, caffeinated, decaffeinated) |
| | Soft drinks, other flavored (artificially sweetened, caffeinated, decaffeinated) |
| | Water, carbonated (artificially sweetened) |
| Cola | Soft drinks, cola types (sugar-sweetened, artificially sweetened, caffeinated, decaffeinated) |
| Tea (sugar-sweetened) | Tea, regular and decaffeinated, hot and cold (sugar-sweetened) |
| Tea (unsweetened) | Tea, regular and decaffeinated, hot and cold (unsweetened) |

**Supplementary Table S2.** Food group mapping of 60-item food frequency questionnaire (FFQ) administered to 1,079 NHANES III participants included in the case-control study of dietary characteristics and monoclonal gammopathy of undetermined significance in the National Health and Nutrition Examination Survey (NHANES)

| Primary FFQ Group | FFQ Item |
|---|---|
| Milk, yogurt, ice cream, cheese | Milk (to drink or on cereal) |
| | Yogurt and frozen yogurt |
| | Chocolate milk and hot cocoa[1] |
| | Ice cream, ice milk, milkshakes[1] |
| | Cheese, all types |
| Meat, poultry, seafood, stews | Beef |
| | Pork and ham |
| | Chicken and turkey |
| | Bacon/sausage/processed meats |
| | Liver and other organ meats |
| | Shrimp, clams, etc. |
| | Fish |
| | Stew or soup with vegetables |
| | Cheese dishes |
| Eggs | Eggs |
| Beans, peanuts | Beans, lentils, chickpeas |
| | Peanuts, peanut butter, etc. |
| Bread, cereal, rice, starchy entrees, cakes | White bread, rolls, etc. |
| | Dark breads and rolls |
| | Corn bread, muffins, tortillas |
| | Flour tortillas |
| | Cakes, cookies, brownies, etc. [1] |
| | Salted snacks |
| | Cooked, hot cereals |

| | |
|---|---|
| | Rice |
| | Cereals: All-Bran, etc. |
| | Cereals: Total, etc. |
| | All other cold cereals |
| | Pizza, calzone, lasagna |
| | Spaghetti/pasta w/ tomato sauce |
| Fruits, excluding juices | Citrus fruits |
| | Melons |
| | Peaches, nectarines, etc. |
| | Any other fruits |
| Fruit juices | Orange juice, etc. |
| | Other fruit juices |
| Vegetables | White potatoes |
| | Broccoli |
| | Spinach, greens, etc. |
| | Carrots |
| | Sweet potatoes, yams, etc. |
| | Tomatoes |
| | Brussels sprouts/cauliflower |
| | Tossed salad |
| | Cabbage, coleslaw, sauerkraut |
| | Hot red chili peppers |
| | Other peppers |
| | Any other vegetables |
| Added fats | Margarine |
| | Butter |
| | Oil/vinegar, mayonnaise, etc. |

| | |
|---|---|
| Desserts, candy, beverages | Chocolate candy and fudge |
| | Chocolate milk and hot cocoa[1] |
| | Ice cream, ice milk, milkshakes[1] |
| | Cakes, cookies, brownies, etc.[1] |
| | Hi-C, Tang, Koolaid, etc. |
| | Diet colas, diet sodas, etc. |
| | Regular colas and sodas |
| | Regular coffee |
| | Regular tea |
| | Beer and lite beer |
| | Wine, etc. |
| | Hard liquor |

FFQ=food frequency questionnaire. [1]For comparability with the USDA Food Coding Scheme used in the 24-hour dietary recall, "chocolate milk and hot cocoa" and "ice cream, ice milk, and milkshakes" are assigned to the "Milk, yogurt, ice cream, and cheese" group but for the purposes of this analysis are also shown with the "Desserts, candy, and beverages" group. For USDA comparability, "cakes, cookies, brownies, etc." are assigned to "Bread, cereal, rice, starchy entrees, cakes" but are also grouped with "Desserts, candy, and beverages" group for this analysis.

| **Supplementary Table S3.** Custom food groups constructed from 60-item food frequency questionnaire (FFQ) administered to 1,079 NHANES III participants included in the case-control study of dietary characteristics and monoclonal gammopathy of undetermined significance in the National Health and Nutrition Examination Survey (NHANES) | |
|---|---|
| **Custom FFQ Food Group** | **FFQ Item** |
| Processed meat | Bacon/sausage/processed meats |
| Tomatoes | Tomatoes |
| Cruciferous vegetables | Broccoli |
| | Brussels sprouts/cauliflower |
| | Cabbage, coleslaw, sauerkraut |
| Sugar-sweetened foods and beverages | Ice cream, ice milk, milkshakes |
| | Cakes, cookies, brownies, etc. |
| | Chocolate candy and fudge |
| | Hi-C, Tang, Koolaid, etc. |
| | Regular colas and sodas |
| Sugar-sweetened beverages | Hi-C, Tang, Koolaid, etc. |
| | Regular colas and sodas |
| Sugar-sweetened beverages plus juices | Hi-C, Tang, Koolaid, etc. |
| | Regular colas and sodas |
| | Orange juice, etc |
| | Other fruit juices |
| Soft drinks | Diet colas, diet sodas, etc. |
| | Regular colas and sodas |
| Soft drinks (sugar-sweetened) | Regular colas and sodas |
| Soft drinks (artificially sweetened) | Diet colas, diet sodas, etc. |
| Tea | Regular tea |

**Supplementary Table S4.** Association between monthly food intake* by primary food frequency questionnaire group and overall, low-risk, and intermediate/high-risk MGUS in 1,079 NHANES III participants in the case-control study of dietary characteristics and monoclonal gammopathy of undetermined significance in the National Health and Nutrition Examination Survey (NHANES)

| | **n (%)** | | | | **OR (95% CI)[1,2,3]** | | |
|---|---|---|---|---|---|---|---|
| | **MGUS** | **LRMGUS** | **IHRMGUS** | **Non-MGUS** | **MGUS** | **LRMGUS** | **IHRMGUS** |
| **Primary FFQ Group, times per month** | | | | | | | |
| Milk, yogurt, ice cream, cheese [4] | | | | | | | |
| T1 (0-25) | 75 (33.8) | 29 (29.0) | 40 (38.8) | 280 (32.8) | ref. | ref. | ref. |
| T2 (26-44) | 82 (36.9) | 36 (36.0) | 39 (37.9) | 285 (33.4) | 1.08 (0.75-1.55) | 1.19 (0.70-2.02) | 0.95 (0.58-1.55) |
| T3 (45-187) | 65 (29.3) | 35 (35.0) | 24 (23.3) | 289 (33.8) | 0.84 (0.57-1.25) | 1.11 (0.63-1.93) | 0.59 (0.33-1.04) |
| Meat, poultry, seafood, stews | | | | | | | |
| T1 (0-31) | 50 (22.6) | 23 (23.0) | 24 (23.5) | 275 (32.3) | ref.** | ref. | ref. |
| T2 (32-48) | 86 (38.9) | 36 (36.0) | 41 (40.2) | 292 (34.3) | **1.64 (1.11-2.42)** | 1.45 (0.83-2.52) | 1.66 (0.97-2.83) |
| T3 (49-221) | 85 (38.5) | 41 (41.1) | 37 (36.3) | 285 (33.5) | **1.70 (1.14-2.52)** | 1.71 (0.98-2.96) | 1.56 (0.90-2.71) |
| Eggs | | | | | | | |
| T1 (0-2) | 76 (34.2) | 31 (31.0) | 37 (35.9) | 301 (35.1) | ref. | ref. | ref. |
| T2 (3-9) | 80 (36.0) | 40 (40.0) | 34 (33.0) | 327 (38.2) | 0.98 (0.69-1.40) | 1.19 (0.72-1.97) | 0.87 (0.53-1.44) |
| T3 (10-61) | 66 (29.7) | 29 (29.0) | 32 (31.1) | 229 (26.7) | 1.17 (0.79-1.72) | 1.29 (0.74-2.27) | 1.10 (0.64-1.88) |
| Beans, peanuts | | | | | | | |
| T1 (0-4) | 69 (31.1) | 29 (29.0) | 35 (34.0) | 303 (35.4) | ref. | ref. | ref. |
| T2 (5-13) | 78 (35.1) | 34 (34.0) | 37 (35.9) | 273 (31.9) | 1.26 (0.87-1.81) | 1.30 (0.77-2.20) | 1.18 (0.72-1.93) |
| T3 (14-161) | 75 (33.8) | 37 (37.0) | 31 (30.1) | 281 (32.8) | 1.19 (0.81-1.75) | 1.46 (0.85-2.49) | 0.94 (0.54-1.61) |
| Bread, cereal, rice, starchy entrees, cakes [5] | | | | | | | |
| T1 (0-69) | 82 (37.1) | 40 (40.4) | 33 (32.0) | 284 (33.5) | ref. | ref. | ref. |
| T2 (70-102) | 67 (30.3) | 27 (27.3) | 38 (36.9) | 281 (33.2) | 0.83 (0.57-1.21) | 0.67 (0.40-1.13) | 1.18 (0.91-1.96) |
| T3 (103-385) | 72 (32.6) | 32 (32.3) | 32 (31.1) | 282 (33.3) | 0.88 (0.61-1.28) | 0.79 (0.47-1.32) | 0.98 (0.58-1.67) |
| Fruits, excluding juice, and vegetables | | | | | | | |
| T1 (0-74) | 64 (28.8) | 32 (32.0) | 27 (26.2) | 283 (33.3) | ref. | ref. | ref. |
| T2 (75-127) | 86 (38.7) | 42 (42.0) | 39 (37.9) | 284 (33.4) | 1.36 (0.94-1.97) | 1.30 (0.79-2.15) | 1.47 (0.87-2.49) |
| T3 (128-375) | 72 (32.4) | 26 (26.0) | 37 (35.9) | 284 (33.4) | 1.13 (0.77-1.68) | 0.80 (0.45-1.41) | 1.42 (0.82-2.47) |
| Fruits, excluding juice | | | | | | | |
| T1 (0-14) | 64 (28.8) | 26 (26.0) | 32 (31.1) | 286 (33.5) | ref. | ref. | ref. |
| T2 (15-35) | 70 (31.5) | 41 (41.0) | 26 (25.2) | 281 (32.9) | 1.13 (0.77-1.65) | 1.68 (0.99-2.84) | 0.80 (0.46-1.39) |
| T3 (36-183) | 88 (39.6) | 33 (33.0) | 45 (43.7) | 288 (33.7) | 1.37 (0.95-1.99) | 1.30 (0.75-2.25) | 1.39 (0.85-2.29) |

| | | | | | | | |
|---|---|---|---|---|---|---|---|
| Vegetables | | | | | | | |
| T1 (0-51) | 67 (30.2) | 34 (34.0) | 27 (26.2) | 287 (33.7) | ref. | ref. | ref. |
| T2 (52-90) | 70 (31.5) | 31 (31.0) | 36 (35.0) | 284 (33.3) | 1.07 (0.73-1.56) | 0.91 (0.54-1.53) | 1.39 (0.82-2.36) |
| T3 (91-307) | 85 (38.3) | 35 (35.0) | 40 (38.8) | 282 (33.1) | 1.33 (0.91-1.94) | 1.05 (0.62-1.77) | 1.60 (0.93-2.75) |
| Fruit juice | | | | | | | |
| T1 (0-7) | 50 (22.5) | 26 (26.0) | 19 (18.5) | 277 (32.4) | ref.** | ref. | ref.** |
| T2 (8-30) | 103 (46.4) | 48 (48.0) | 45 (43.7) | 372 (43.5) | **1.54 (1.06-2.25)** | 1.38 (0.83-2.29) | **1.80 (1.02-3.16)** |
| T3 (31-244) | 69 (31.1) | 26 (26.0) | 39 (37.9) | 206 (24.1) | **1.85 (1.23-2.79)** | 1.36 (0.76-2.44) | **2.77 (1.54-4.98)** |
| Added fats | | | | | | | |
| T1 (0-17) | 68 (30.8) | 27 (27.0) | 36 (35.3) | 279 (32.8) | ref. | ref. | ref. |
| T2 (18-38) | 64 (29.0) | 36 (36.0) | 22 (21.6) | 266 (31.2) | 1.00 (0.68-1.46) | 1.40 (0.82-2.38) | 0.64 (0.37-1.13) |
| T3 (39-215) | 89 (40.3) | 37 (37.0) | 44 (43.1) | 307 (36.0) | 1.23 (0.85-1.79) | 1.24 (0.72-2.13) | 1.18 (0.72-1.94) |
| Desserts, candy, beverages [4,5] | | | | | | | |
| T1 (0-60) | 64 (29.0) | 28 (28.0) | 31 (30.4) | 288 (33.8) | ref. | ref. | ref. |
| T2 (61-109) | 95 (43.0) | 46 (46.0) | 39 (38.2) | 283 (33.2) | **1.56 (1.08-2.24)** | 1.59 (0.96-2.64) | 1.42 (0.85-2.37) |
| T3 (110-727) | 62 (28.1) | 26 (26.0) | 32 (31.4) | 281 (33.0) | 1.03 (0.69-1.54) | 0.87 (0.49-1.55) | 1.22 (0.71-2.10) |

*Intake estimated by 60-item food frequency questionnaire (FFQ). MGUS=monoclonal gammopathy of undetermined significance; LRMGUS=low-risk MGUS; IHRMGUS=intermediate/high-risk MGUS; OR=odds ratio; CI=confidence interval; n=number; T=tertile. [1] Adjusted for age, sex, race, body mass index, and total daily energy intake. [2] Significance of overall category based on p-for-trend. **p for trend < 0.05. [3] Odds of MGUS, LRMGUS, and IHRMGUS all relative to non-MGUS. [4] Chocolate milk and ice cream are included in both the “Milk, yogurt, ice cream, cheese” and the “Desserts, candy, beverages” groups. [5] Cakes, cookies, brownies are included in both the “Bread, cereal, rice, starchy entrees, cakes” and “Desserts, candy, beverages” groups. **Bold text** indicates that n-tile is statistically significant.

**Supplementary Table S5.** Association between monthly food intake* of select custom food frequency questionnaire food groups and overall, low-risk, and intermediate/high-risk MGUS in 1,079 NHANES III participants in the case-control study of dietary characteristics and monoclonal gammopathy of undetermined significance in the National Health and Nutrition Examination Survey (NHANES)

| | n (%) | | | | OR (95% CI)[1,2] | | |
|---|---|---|---|---|---|---|---|
| | MGUS | LRMGUS | IHRMGUS | Non-MGUS | MGUS | LRMGUS | IHRMGUS |
| **FFQ Group, times per month** | | | | | | | |
| Processed meat | | | | | | | |
| T1 (0-1) | 54 (24.3) | 23 (23.0) | 25 (24.3) | 255 (29.8) | ref. | ref. | ref. |
| T2 (2-8) | 77 (34.7) | 30 (30.0) | 41 (39.8) | 273 (31.9) | 1.35 (0.91-1.99) | 1.21 (0.68-2.15) | 1.57 (0.92-2.67) |
| T3 (9-75) | 91 (41.0) | 47 (47.0) | 37 (35.9) | 329 (38.4) | 1.35 (0.92-1.99) | 1.60 (0.93-2.75) | 1.15 (0.66-2.00) |
| Tomatoes | | | | | | | |
| T1 (0-4) | 88 (39.6) | 42 (42.0) | 39 (37.9) | 337 (39.3) | ref. | ref. | ref. |
| T2 (5-13) | 62 (27.9) | 26 (26.0) | 33 (32.0) | 253 (29.5) | 0.93 (0.65-1.35) | 0.82 (0.49-1.38) | 1.13 (0.69-1.86) |
| T3 (14-91) | 72 (32.4) | 32 (32.0) | 31 (30.1) | 267 (31.2) | 1.04 (0.72-1.50) | 0.96 (0.58-1.59) | 1.03 (0.61-1.75) |
| Cruciferous vegetables | | | | | | | |
| T1 (0-3) | 64 (28.8) | 31 (31.0) | 29 (28.2) | 254 (29.7) | ref. | ref. | ref. |
| T2 (4-9) | 73 (32.9) | 34 (34.0) | 35 (34.0) | 305 (35.6) | 0.95 (0.65-1.38) | 0.90 (0.53-1.50) | 1.02 (0.61-1.72) |
| T3 (10-134) | 85 (38.3) | 35 (35.0) | 39 (37.9) | 297 (34.7) | 1.12 (0.78-1.63) | 0.93 (0.55-1.56) | 1.16 (0.69-1.94) |
| Sugar-sweetened foods and beverages | | | | | | | |
| T1 (0-15) | 72 (32.4) | 27 (27.0) | 42 (40.8) | 288 (33.6) | ref. | ref. | ref. |
| T2 (16-40) | 76 (34.2) | 37 (37.0) | 28 (27.2) | 279 (32.6) | 1.09 (0.76-1.58) | 1.35 (0.79-2.30) | 0.71 (0.42-1.19) |
| T3 (41-343) | 74 (33.3) | 36 (36.0) | 33 (32.0) | 289 (33.8) | 1.05 (0.72-1.53) | 1.27 (0.73-2.19) | 0.82 (0.49-1.35) |
| Sugar-sweetened beverages | | | | | | | |
| None | 77 (34.7) | 38 (38.0) | 34 (33.0) | 315 (36.8) | ref. | ref. | ref. |
| Q1 (1-13) | 77 (34.7) | 35 (35.0) | 34 (33.0) | 271 (31.6) | 1.16 (0.81-1.66) | 1.06 (0.65-1.74) | 1.18 (0.71-1.95) |
| Q2 (14-304) | 68 (30.6) | 27 (27.0) | 35 (34.0) | 271 (31.6) | 1.03 (0.70-1.53) | 0.76 (0.43-1.32) | 1.30 (0.76-2.21) |
| Soft drinks | | | | | | | |
| T1 (0-3) | 72 (32.4) | 29 (29.0) | 39 (37.9) | 292 (34.1) | ref. | ref. | ref. |
| T2 (4-16) | 71 (32.0) | 36 (36.0) | 27 (26.2) | 267 (31.2) | 1.09 (0.75-1.59) | 1.37 (0.80-2.32) | 0.76 (0.45-1.30) |
| T3 (17-304) | 79 (35.6) | 35 (35.0) | 37 (35.9) | 297 (34.7) | 1.13 (0.77-1.65) | 1.14 (0.65-1.97) | 1.03 (0.62-1.72) |
| Soft drinks (sugar-sweetened) | | | | | | | |
| None | 108 (48.7) | 52 (52.0) | 51 (49.5) | 405 (47.3) | ref. | ref. | ref. |
| Q1 (1-9) | 61 (27.5) | 27 (27.0) | 26 (25.2) | 221 (25.8) | 1.02 (0.72-1.47) | 0.92 (0.56-1.52) | 0.95 (0.57-1.57) |
| Q2 (10-304) | 53 (23.9) | 21 (21.0) | 26 (25.2) | 231 (27.0) | 0.86 (0.59-1.27) | 0.62 (0.35-1.10) | 0.98 (0.58-1.66) |

| | | | | | | | |
|---|---|---|---|---|---|---|---|
| Soft drinks (artificially sweetened) | | | | | | | |
| None | 148 (66.7) | 66 (66.0) | 69 (67.0) | 587 (68.6) | ref. ** | ref. ** | ref. |
| Q1 (1-12) | 24 (10.8) | 9 (9.0) | 13 (12.6) | 129 (15.1) | 0.74 (0.46-1.19) | 0.65 (0.31-1.35) | 0.84 (0.45-1.58) |
| Q2 (13-182) | 50 (22.5) | 25 (25.0) | 21 (20.4) | 140 (16.4) | **1.48 (1.01-2.18)** | 1.67 (0.99-2.81) | 1.32 (0.76-2.28) |
| Tea | | | | | | | |
| None | 127 (57.2) | 63 (63.0) | 54 (52.4) | 444 (51.8) | ref. | ref. | ref. |
| Q1 (1-12) | 44 (19.8) | 19 (19.0) | 22 (21.4) | 192 (22.4) | 0.80 (0.54-1.18) | 0.66 (0.38-1.15) | 0.96 (0.56-1.63) |
| Q2 (13-243) | 51 (23.0) | 18 (18.0) | 27 (26.2) | 221 (25.8) | 0.81 (0.56-1.16) | **0.55 (0.32-0.96)** | 1.03 (0.62-1.69) |

*Intake estimated by 60-item food frequency questionnaire (FFQ). MGUS=monoclonal gammopathy of undetermined significance; LRMGUS=low-risk MGUS; IHRMGUS=intermediate/high-risk MGUS; OR=odds ratio; CI=confidence interval; n=number; T=tertile; Q=quantile, used when tertiles are not practical. [1] Adjusted for age, sex, race, body mass index, and total daily energy intake. [2] Odds of MGUS, LRMGUS, and IHRMGUS all relative to non-MGUS. **Bold text** indicates that n-tile is statistically significant. **p for trend < 0.05.
